# COVID-19 cases associated with testing policies in 34 highly democratic countries, 2020-2022

**DOI:** 10.1101/2023.10.19.23297270

**Authors:** Leon S. Robertson

**Author notes:** The data in this study were obtained from public sources and did not involve contacts with individuals who would require informed consent. Therefore, no ethical board committee was involved. The research was not funded. The author has no financial or other interests that would be affected by the publication of this paper.

## Abstract

COVID-19 testing policies varied in time from testing only the symptomatic, testing the symptomatic and persons at higher risk of severe disease, on-demand testing for people who wanted one, and two periods of government-imposed mass testing in Slovakia. Using Poisson regression, this study examines the associations of COVID-19 cases during the times that the noted policies were in effect in 34 countries rated highest on democracy scores. Statistically corrected for other risk factors, increases in negative tests were associated with subsequent surges in cases when on-demand testing was promoted, particularly when coupled with poor contact tracing. Mass testing in Slovakia was associated with reduced spread of the virus for short periods but was deemed unsustainable. The data support the hypothesis that on-demand testing resulted in the unanticipated consequence of increased travel and increased exposure of travelers to the virus.

## Introduction

When tests for the SARS-COV-2 virus became available, the decision of whom to test varied among countries and jurisdictions within countries. When it became known that asymptomatic carriers of the virus could transmit it to others, governments extended diagnostic testing of the symptomatic to asymptomatic people at higher risk of severe disease and death and their caregivers in many jurisdictions. During some periods, governments promoted on-demand testing at drive-in sites and other venues or by appointment. The objective of on-demand testing was to identify the positives and trace and test their contacts, recommending or requiring those who tested positive to isolate themselves for specified times. The percent testing positive was considered an indicator of when restrictions to protect public health should be adjusted. Slovakia implemented a plan to test everyone in a large segment of the population using antigen tests that provide quick results but with less accuracy than polymerase chain reaction (PCR) tests that take 24 or more hours to obtain results [1]. The objective was to require those who tested positive as well as those untested to isolate themselves. In this case, since all positives, tested or not, would presumably be isolated, minimizing transmission, there was no contact tracing. In a later 3- month period of attenuated mass testing, vigorous contact tracing was employed.

During the first seven months of the COVID-19 pandemic in 2020, Slovakia substantially contained the spread of SARS-CoV-2. The government screened and quarantined infected travelers from abroad, closed nonessential businesses, required mask use, and limited the size of gatherings. A four-phase plan to reduce restrictions was adopted and was implemented as spring ended. When a surge in cases occurred in October many of the restrictions were reimposed [1].

In October, Slovakia conducted a pilot study of mass testing using rapid antigen tests in four counties that were most affected followed by a plan to test all persons 10 to 65 years old in the country with some exceptions [2]. On the weekend of 31 October – 1 November 2020, the government implemented the plan. The following weekend the effort was repeated in counties that had test positivity above .07 percent. In a population of about 5.4 million persons more than 3.6 million people and 2.0 million people respectively were tested during the two autumn weekends. People who were not tested were required to stay at home for 10 days with a potential fine of 1650 Euros for non-compliance [2].

A team of researchers studied the changes in cases from the first to the second weekend of the autumn mass testing and reported a reduction of about 60 percent. Based on a simulation of restrictions and testing results in one county, they concluded that mass testing was mostly associated with reduced cases when combined with the other countermeasures. They appropriately cautioned, “The observational nature of this study made it difficult to clearly distinguish the effect of the mass testing campaign from that of other non-pharmaceutical interventions introduced at a similar time, that have led to a reduction of contacts and mobility albeit much less than during the spring lockdown” [3].

Testing advocates ignored the caution. One pair of authors cited a preprint of the Slovakian study as a rationale for unlimited testing. “Multiple countries have been successful at controlling SARS-CoV-2 transmission by investing in large-scale testing capacity” [4] they wrote, also citing an article regarding New Zealand that said the opposite. The New Zealand authors wrote that extensive lockdown procedures were adopted in New Zealand because “the country didn’t have sufficient testing and contact-tracing capacity to contain the virus” [5]. At the end of 2020, New Zealand’s cumulated testing rate per population was about 36 percent that of the U.S., and its cumulated case rate per million population was 348 compared to 57,872 in the U.S. [6]. New Zealand had among the lowest case rates in the world while testing only the symptomatic and vulnerable until mid-December 2021 when it changed to distributing free home test kits after which its cases increased. [7]

Another study of the autumn 2020 mass testing in Slovakia compared regions of the country and concluded that there was a reduction in cases but the effort might have been more successful if the testing had been confined to the regions that had more cases. The authors noted that people who spent an hour or more at the testing sites for tests and results had a high probability of exposure to an infected person but did not account for the possibility that a negative test could result in increased travel and further exposure. [8]

In 2021 Slovakia tested 200,000-350,000 people per day for people who wanted to avoid a stay-at-home order from 18 January to late April. People who lived in high-risk areas were required to get a test weekly. Effective 27 January, persons attending a variety of venues had to show evidence of negative test results to be admitted. People with certain illnesses, children less than 10 years old or in certain schools, and the vaccinated were exempt [9]. A third study of the autumn testing effort compared counties matched by various criteria and concluded that the reduction in transmissibility and cases resulting from mass testing in autumn was probably no higher than 30 percent. The authors judged the data “too noisy” to assess the consequences of the winter effort [10].

On-demand testing is different from mass testing imposed by the government. Research on the consequences of on-demand testing among and within countries in 2020 found that increases in negative tests predicted surges in COVID-19 hospitalizations 14 days later while limiting tests to the symptomatic and vulnerable was associated with fewer hospitalizations [11]. Self-selection to be tested is unlikely to yield a representative sample of the population. When tests are offered on demand, often free, most of the persons seeking tests are doing so intending to visit or travel rather than because of symptoms or contact with the infected. In the U.S. each negative test was followed by an average of 334 kilometers of road vehicle travel the week after the test in the spring and summer of 2020. COVID-19 cases per population among U.S. counties were strongly predicted by road travel [7]. If the travelers came in close contact with infected persons, they were likely to become infected.

In the study reported here, the associations of COVID-19 cases with negative tests, positivity, contact tracing, and the delta and omicron variants are included to control statistically for the associations with these factors. The data were analyzed separately for periods of on-demand testing, testing the symptomatic and vulnerable, and testing only the symptomatic. Data from Slovakia was analyzed separately to account for mass testing during the noted periods there. The study was confined to 34 of 36 strongly democratic countries because less democratic governmental regimes have been suspected of underreporting cases. Two of the countries that contained the virus most successfully, Iceland and New Zealand [7], were excluded because they did not report positivity.

## Methods

Daily data on COVID-19 cases, tests, and test positivity through December 2022 were downloaded on 9 July 2023 from ourworldindata.org [12]. Negative tests were derived by multiplying positivity by total tests and subtracting the positives from the total. Daily ratings of country efforts at contact tracing were downloaded on 9 July 2023 from a group at Oxford University. The data are coded as 0=no tracing, 1=limited cases, 2=comprehensive contact tracing [13]. The study includes all but the mentioned two countries that had a democracy score above .80 on a scale of zero to 1 based on a variety of freedoms, rights, and equal treatment of citizens. The scores range from .811 (U.S.) to .958 (Denmark) [14,15].

Because of weekday fluctuations in reporting, the variables were averaged over seven days. To reduce skew in the frequency distributions, the logarithms of the continuously distributed predictor variables were used. Poison regression was employed to estimate the association of the predictor variables to average daily cases with the logarithm of the population of a given country as the offset variable. Where t=time in days, the regression model is:

Cases_t_= a + b1 log(cases_t-14_) +b2 log(average negative tests_t-14_) +b3 log(average percent tests positive_t-14_) + b4 (contact tracing) + b5 (delta variant prevalent_t-14_) + b6 (omicron variant prevalent_t-14_) + b7 (t).

The 14-day lag time allowed for travel after testing and incubation of new infections, as used in previous research [7,11]. Since the variables are 7-day averages, the 14-day lag in cases is the average cases 8-14 days after the measured predictors. Controlling statistically for previous cases adjusts for the status of the spread at a given time.

The days from 1 June 2021, to the end of the year were assigned one, otherwise zero to control statistically for the more easily spread delta variant. The days in 2022 through December were assigned one to account for the omicron variant prevalence. Most countries varied their testing policies from time to time so the analysis was done separately for days of on-demand testing, days testing the symptomatic and persons more vulnerable to severe consequences, and days testing only the symptomatic [16]. In a separate analysis of the data from Slovakia, a lagged binary variable (1 for mass testing days, otherwise zero) was added to the equation for the autumn mass testing days rather than include the number of tests conducted on those days. The number of tests on the autumn mass testing days was not included in the data from ourworldindata.org and, if it had been, would have skewed the frequency distributions beyond correction by logarithms. A separate binary variable was used to estimate the association of cases with the winter-spring testing effort. Except for the mass testing dates, Slovakia allowed on-demand testing during some periods but tested only the symptomatic during others. The mass testing was done during periods when only the symptomatic were being otherwise tested.

## Results

Figure 1. shows the cases per population per day for each of the testing policies subdivided by the best versus lower ratings on contact tracing. When on-demand testing was allowed or encouraged and contact tracing was poorly executed or nonexistent, the case rate was about twice that when testing was limited to the symptomatic and vulnerable and 4-5 times the rate when the symptomatic only were tested. When contact tracing was comprehensive during on-demand testing periods, the rate was similar to the rate during symptomatic and vulnerable testing with poor tracing and about twice that when the symptomatic only were tested with comprehensive contact tracing.

**Figure 1.**
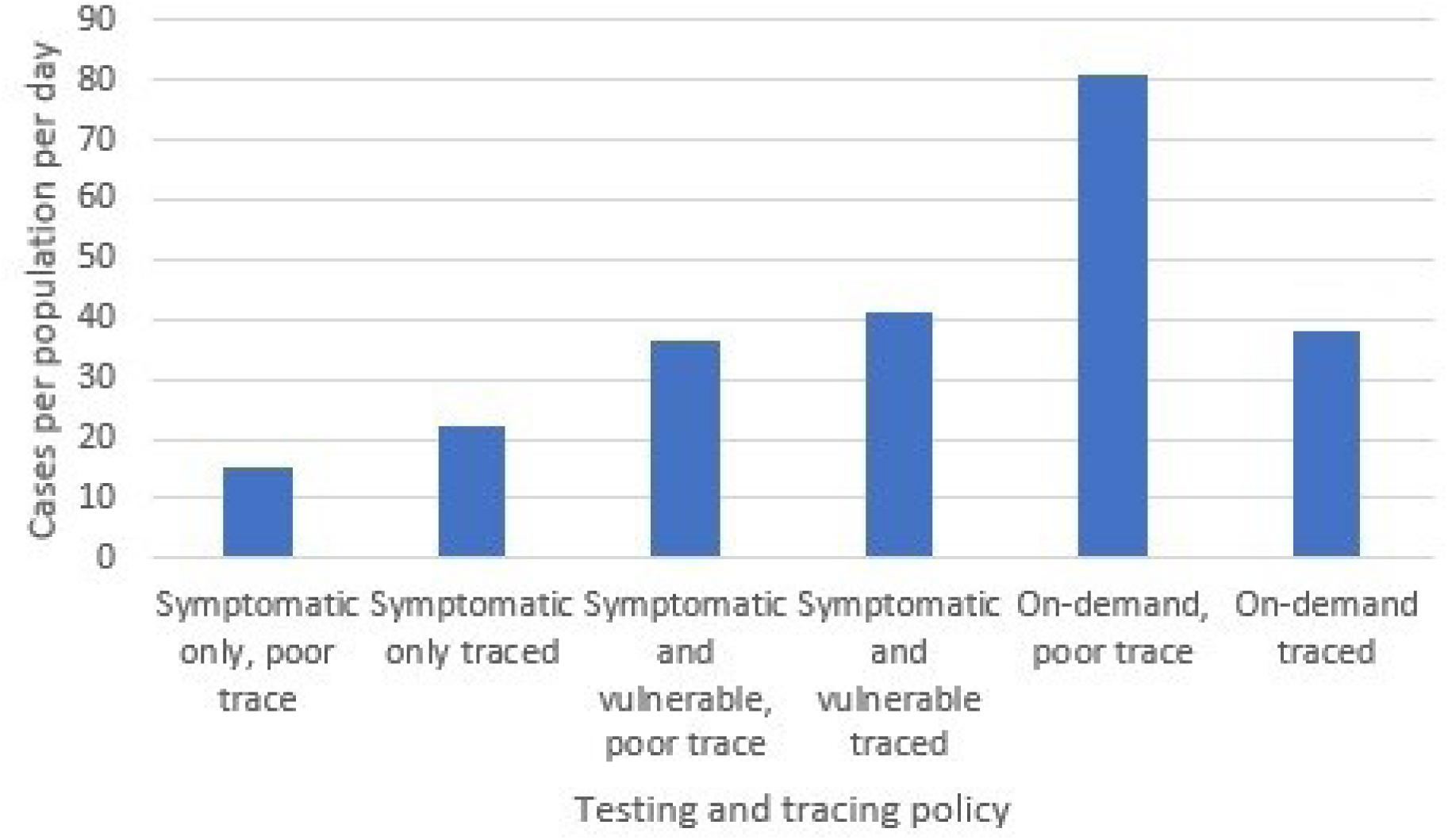
Cases per 100,000 population per day by testing policy and contact tracing rating, 32 democratic countries, 2020-2022.

Odds ratios based on the regression coefficients and confidence intervals are in Table 1. Corrected statistically for the other predictors, surges in COVID-19 cases were predicted by increased negative tests when testing was on demand and when only the symptomatic were tested but not when testing was limited to the symptomatic and vulnerable. The same was found for the association with positivity. Good contact tracing was associated with reduced cases when the policy was test-on-demand or the symptomatic and vulnerable were tested but not when only the symptomatic were tested. Adjusted for these factors, the cases declined in time when on-demand testing was allowed but increased in time during symptomatic testing only or including the vulnerable. In Slovakia, the autumn 2020 mass testing was associated with about 27 percent lower odds of cases per day, and the winter-spring 2021 testing was associated with about a 35 percent decrease in odds of infection per day. The latter during about three months means that the virus was mainly contained by the end of April 2021. Although Slovakia vigorously traced the contacts of those who tested positive until May 2021, the contact tracing rating was reduced from comprehensive to limited after that, and the delta and omicron variants later produced large surges in cases.

**Table 1.** Poisson regression-based odds ratios and 95 percent confidence intervals of variables hypothesized to predict daily COVID-19 cases in Slovakia and 33 other democratic countries by testing policy, 2020-2022.

| Predictor variable | Test on demand | Test symptomatic and vulnerable | Only test symptomatic | Slovakia |
| --- | --- | --- | --- | --- |
| Log(cases before) | .764<br>(.763, .765) | 1.690<br>(1.689, 1.691) | .895<br>(.894, .896) | 2.880<br>(2.856, 2.904) |
| Log(negative tests) | 1.728<br>(1.726, 1.730) | .996<br>(.995, .997) | 2.206<br>(2.201, 2.211) | 1.062<br>(1.054, 1.070) |
| Log(percent tests positive) | 2.832<br>(2.829, 2.835) | .966<br>(.965, .967) | 2.588<br>(2.583, .2.563) | .787<br>(.780, .794) |
| Contact tracing | .633<br>(.633, .633) | .662<br>(.662, .662) | 1.435<br>(1.433, 1.437) | 2.067<br>(1.958, 2.170) |
| Delta variant | 2.289<br>(2.286, 2.292) | 1.206<br>(1.204, 1.208) | 1.271<br>(1.269, 1.273) | 3.827<br>(3.636, 4.026) |
| Omicron variant | 4.665<br>(4.660, 4.670) | 2.257<br>(2.255, 2.259) | 2.185<br>(2.181, 2.189) | 4.768<br>(4.527, 5.023) |
| Log(time) | .304<br>(.303, .305) | 1.466<br>(1.465, 1.467) | 1.208<br>(1.206, 1.210) | .473<br>(.466,.480) |
| Autumn mass testing |  |  |  | .734<br>(.715, .756) |
| Winter-Spring Testing |  |  |  | .644<br>(.635, .673) |
| N=days | 10305 | 7689 | 6375 | 834 |

## Discussion

The finding that surges in negative tests predict subsequent increased infections during on-demand testing periods is consistent with previous research [7,11]. This study indicates that this is particularly so when the contact tracing system is nonexistent or poorly executed. In the U.S. contact tracers were overwhelmed. On about 70 percent of days in U.S. states, there were more cases than contact tracers had time to trace [11]. Even when tracing contacts was considered adequate, failure to quarantine by those who tested positive may have been a factor in the spread of the virus. A study of contact tracing in the U.S. found a substantial lack of cooperation in identifying contacts and self-quarantine [17]. Another U.S. study claimed that testing and contact tracing were effective [18] in selected districts but did not account for the cases that likely occurred due to travel after negative tests. These would have to be subtracted to assess the net effect of a testing policy. Although the percent positive also predicts surges during periods of on-demand testing, testing individuals is not necessary to predict surges in cases. Wastewater testing has been found to predict surges in a variety of countries [19].

In Slovakia, the association of reduced subsequent cases with mass testing in autumn, 2020 and in winter and spring, 2021 supports the hypothesis that mass testing along with vigorous contact tracing and required evidence of a negative test to travel reduced the spread but the mass testing did not stop the eventual resurgence of the virus. Despite the heroic and apparently effective efforts of late 2020 and early 2021, Slovakia did not repeat the protocol during the surges of the delta and omicron variants and, as of 18 October 2023, had a cumulated death rate per population 3.9 times that of New Zealand [20]. The first author of one of the Slovakian studies was quoted saying that mass testing is not sustainable. In the autumn mass testing period, the logistics required the deployment of about 40,000 military personnel and 20,000 medical personnel who could not neglect other duties for extended periods [21].

Perhaps unanswerable is whether the hype generated in the media^-^ about Slovakia’s mass testing effort [22–25] resulted in the increased volume of on-demand testing in other countries that retained that policy. The proportion of the 34 countries using the policy declined from about two-thirds at times in 2020 to 40 percent in autumn 2021 but the most populated country (U.S.) in the study, which had among the most cases, hospitalizations, and deaths per capita, continued the policy despite evidence that the tracing and testing system could not keep pace with the spread of the virus [11,26–27]. Nevertheless, testing advocates were repeatedly quoted in various media saying that increased testing was the answer to controlling the virus [28–31]. The evidence suggests that the opposite is true particularly when the policy is on-demand testing without good contact tracing.

This study is limited by the use of aggregated data and reliance on correlation to infer causation. Inference of individual behavior from such data can be erroneous and correlation is a necessary but not sufficient condition for inferring causation. The results do pass the plausibility test. If testing was the result of surges in cases, increases in cases would precede increased demand for tests but that is not what happened.

The data strongly suggest that well-intended people urged increased testing and well-intended people sought tests to gain confidence that they were not infected so as not to spread the virus if they traveled [11]. Good intentions are not evidence of effectiveness. Unanticipated consequences of interventions intended to improve health occur frequently enough that results should always be monitored from the time of implementation – witness poison water wells [32], thalidomide babies [33], the opioid epidemic [34], hormone therapy to reduce the symptoms of menopause [35] as examples.

Public health practitioners are faced with a huge dilemma if SARS-CoV-2 should mutate to produce more surges in infections or when the next pandemic resulting from person-to-person transmission arrives, particularly while people remember COVID-19. Free on-demand testing is popular and those who deny its potential harmful effect are likely to advocate it. Given the resentments to mandates developed in the COVID-19 pandemic [36–39], requirements to use masks, physically distance ourselves, and get vaccinated are likely to meet as much or more resistance than occurred during 2020-2022.

Data availability: The data in this study was downloaded from the websites noted in the references.

## Data Availability

The data in this study was downloaded from the websites noted in the references.

